# Single-cell physical phenotyping of mechanically dissociated tissue biopsies for fast diagnostic assessment

**DOI:** 10.1101/2021.11.30.21267075

**Authors:** Despina Soteriou, Markéta Kubánková, Christine Schweitzer, Rocío López-Posadas, Rashmita Pradhan, Oana-Maria Thoma, Andrea-Hermina Györfi, Alexandru-Emil Matei, Maximilian Waldner, Jörg H. W. Distler, Stefan Scheuermann, Jens Langejürgen, Regine Schneider-Stock, Raja Atreya, Markus F. Neurath, Arndt Hartmann, Jochen Guck

## Abstract

Rapid and accurate histopathological diagnosis during surgery is critical for clinical decision-making. The prevalent method of intraoperative consultation pathology is time, labour and cost intensive and requires the expertise of trained pathologists. Here, we present an alternative technique for the rapid, label-free analysis of biopsy samples by sequentially assessing the physical phenotype of singularized, suspended cells in high-throughput. This new diagnostic pipeline combines enzyme-free, mechanical dissociation of tissues with real-time deformability cytometry at measurement rates of 100 – 1,000 cells/sec, and machine learning-based analysis. We show that physical phenotype parameters extracted from brightfield images of single cells can be used to distinguish subpopulations of cells in various tissues, without prior knowledge or the need for molecular markers. Further, we demonstrate the potential of our method for inflammatory bowel disease diagnostics. Using unsupervised dimensionality reduction and logistic regression, we accurately differentiate between healthy and tumorous tissue in both mouse and human biopsy samples. The method delivers results within 30 minutes, laying the groundwork for a fast and marker-free diagnostic pipeline to detect pathological changes in solid biopsies.

## Introduction

Changes in physical properties of cells, such as cell size, shape or deformability, are pivotal to the pathology of some diseases and hold great potential as a diagnostic or prognostic marker ^1,2^. In the last decades, a variety of tools have been developed to examine the mechanical properties of cells, including micropipette aspiration, atomic force microscopy, microbead rheometry and optical tweezers and traps ^3,4^. The field has seen an exponential increase in publications that suggest a strong correlation between cell mechanical phenotype and disease state, including sepsis ^5,6^, malaria ^7^, diabetes ^8^, sickle cell anaemia ^9^ and cancer ^10–12^. Unfortunately, these conventional techniques suffer from low cell throughput and the requirement of deep specialist knowledge for operation, which limits their use as a diagnostic tool. Real-time fluorescence and deformability cytometry (RT-FDC) ^13,14^ is one of several new microfluidic techniques ^10,15–21^ that have overcome these drawbacks, allowing the assessment of physical properties of single cells in a label-free and high-throughput manner, opening a new avenue to clinical diagnostics. RT-FDC is not only fast (with up to 1000 cells analysed per second), but in addition to cell deformability it also provides multidimensional information obtained directly from cell images. The diagnostic potential of RT-FDC has been demonstrated in many human disease conditions ranging from leukaemia to bacterial and viral infections including COVID-19 ^22–26^. However, until now, the applicability of the technique was limited to analysing cultured cells or liquid biopsies from blood or bone marrow.

Solid tissue biopsy is the most common method for characterising malignancy and is fundamental in guiding surgeons during intraoperative and perioperative management of cancer patients. Diagnostic assessment of solid tissue biopsies is commonly delivered through intraoperative consultation pathology which relies on histopathological analysis of frozen biopsy sections ^27^. The conventional workflow of intraoperative diagnosis involves numerous processing steps, staining reagents and the microscopic inspection of tissue slices by experienced pathologists for expert analysis. Moreover, sample preparation is time-, resource- and labour-intensive. Alternative workflows have been proposed ^27^, including stimulated Raman spectroscopy ^28,29^, optical coherence tomography ^30^ and confocal microscopy ^31^, but have not yet been implemented. The necessity of an approach that reduces sample preparation and time to diagnosis is therefore imminent.

Here, we present a rapid label-free diagnostic method for solid tissue biopsies. The approach combines the enzyme-free, mechanical dissociation of tissues using a tissue grinder for the quick and simple isolation of viable single cells ^32,33^ with the sequential assessment of cellular physical phenotypes of thousands of individual cells using RT-FDC. First, we screened a panel of mouse tissues and assessed the cell yield, viability and the feasibility of RT-FDC measurement upon the mechanical dissociation of tissue. We demonstrated the ability to distinguish subpopulations of tissue cells purely based on the image-derived physical parameters without prior knowledge or additional molecular labelling, in contrast to conventional flow cytometry, which relies on multi-colour panels of markers for identifying cells. We also showed that our approach can determine inflammatory changes in tissue, based on the measurement of cell deformability in the microfluidic system. Moreover, we examined frozen and fresh biopsy samples from mouse and human and showed for the first time the potential of RT-FDC to distinguish healthy from cancerous tissues, using principal component analysis (PCA) and machine learning on the multi-dimensional data. The findings demonstrate that assessing the physical phenotype of tissue-derived single cells using RT-FDC is an alternative strategy to detect an inflammatory or malignant state. Our procedure, which can deliver results within 30 minutes, has potential as a diagnostic pipeline to detect pathological changes in biopsies and, more generally, to identify and characterize cell populations in tissues in an unbiased and marker-free manner.

## Results

### Physical phenotyping of single cells obtained by mechanical dissociation of solid tissues

Prior to assessing the physical phenotype of cells, the first challenge faced was the quick extraction of single cells from solid tissues on a time scale of minutes, while aiming for a maximally accurate representation of the heterogeneity of cell subpopulations. For this, we used a tissue grinder (TG); a mechanical dissociation device based on counter rotating rows of grinding teeth (Fig. 1) assembled into a Falcon tube ^33^. The device automatically executes a predefined sequence of alternating cutting and grinding steps to isolate single cells from a solid tissue. In total 10 different murine tissues were processed using either TG or conventional enzymatic protocols for comparison (Supplementary Table 1 and 2). Viability was 70-90% in most tissues; cell yield was similar to enzymatic dissociation and tissue dependent (Supplementary Fig. 1). The key advantage of mechanical dissociation was that the processing time took less than 5 minutes per sample, as opposed to tens of minutes or even several hours for the enzymatic protocols. The speed of the extraction presumably helps to preserve biochemical and biophysical phenotypes in conditions close to those in situ.

**Fig. 1:**
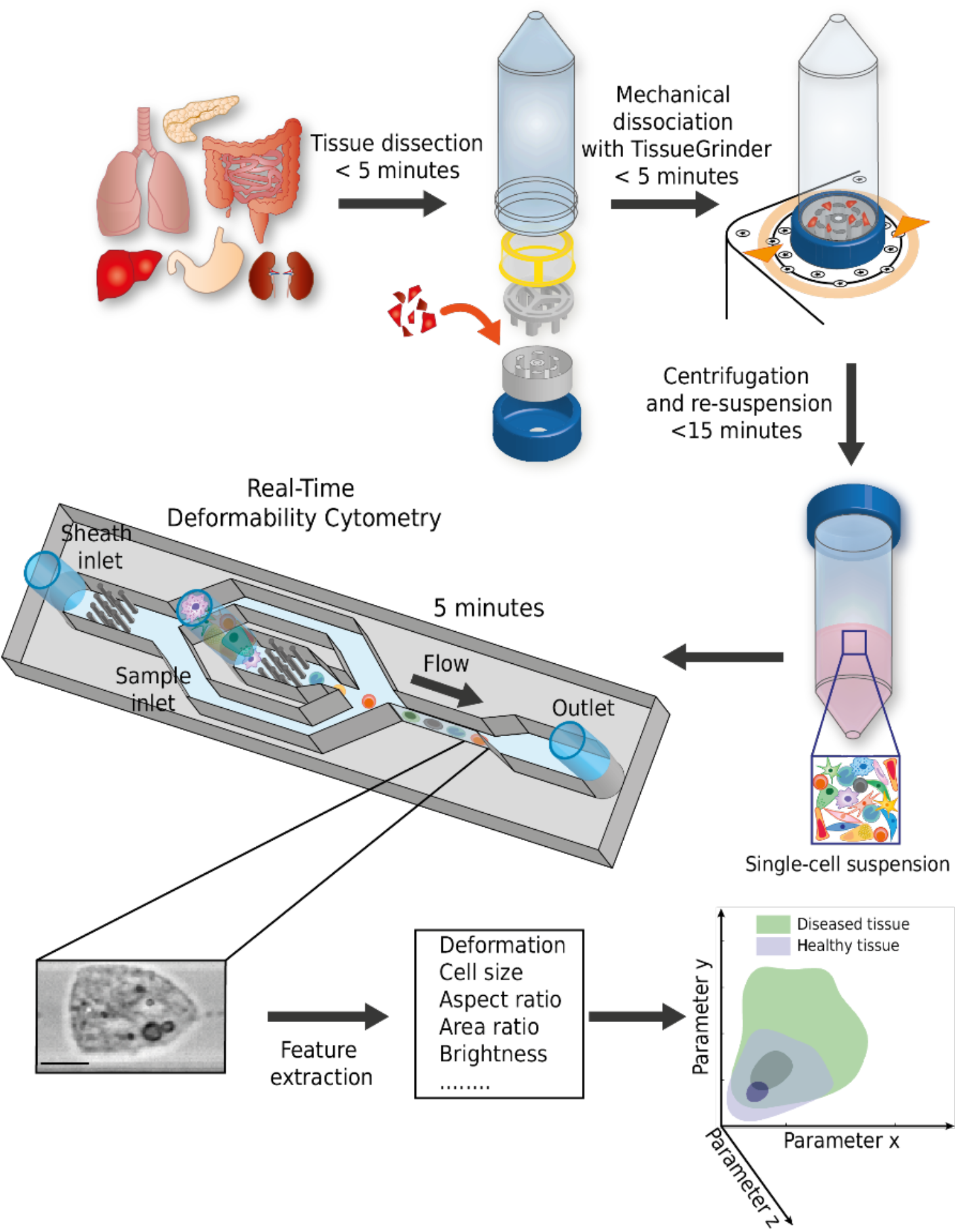
Schematic representation of the analysis procedure. The tissue sample is dissected in small pieces and placed into the inner rotor of the tissue grinder unit containing culture medium. Mechanical dissociation is performed by a pre-programmed, automatically executed sequence of clockwise and anticlockwise rotations. Dissociated cells are centrifuged and resuspended in measuring buffer. The sample is loaded onto a microfluidic chip and analysed using RT-FDC. A brightfield image of every single one of typically 10,000 cells is captured. Various features are extracted from the images, which are used for multi-dimensional analysis. In total, the procedure from tissue to result takes less than 30 minutes.

Next, the extracted single cells were analysed using RT-FDC. In an RT-FDC measurement, hundreds of cells per second, suspended in a high-viscosity medium, are pushed through a microfluidic channel constriction, where they are deformed by shear stress and pressure gradients and an image of each cell is obtained. Several physical parameters were calculated from the images in real time, namely deformation, cell size, brightness, standard deviation of brightness, aspect ratio and area ratio (for details see Supplementary Table 3). Additionally, the fluorescence module ^14^ was used to detect the expression of cell surface markers.

Representative scatter plots of certain physical parameters of cells from liver, colon and kidney are shown in Fig. 2, which revealed distinct clusters of cells. Each of these clusters was composed of cells with similar physical phenotype, typically of the same cell type. This was confirmed by labelling the cells with surface markers for three major cell types: epithelial cells (EpCAM), endothelial cells (CD31), leukocytes (CD45) (Supplementary Fig. 2). For example, a cluster of cells with similar physical properties (in this case defined by average brightness and cell size) was mainly composed of EpCAM positive cells (Fig. 2a), demonstrating that a clean population of epithelial cells can be distinguished in a label-free manner, purely using image-derived physical parameters.

**Fig. 2:**
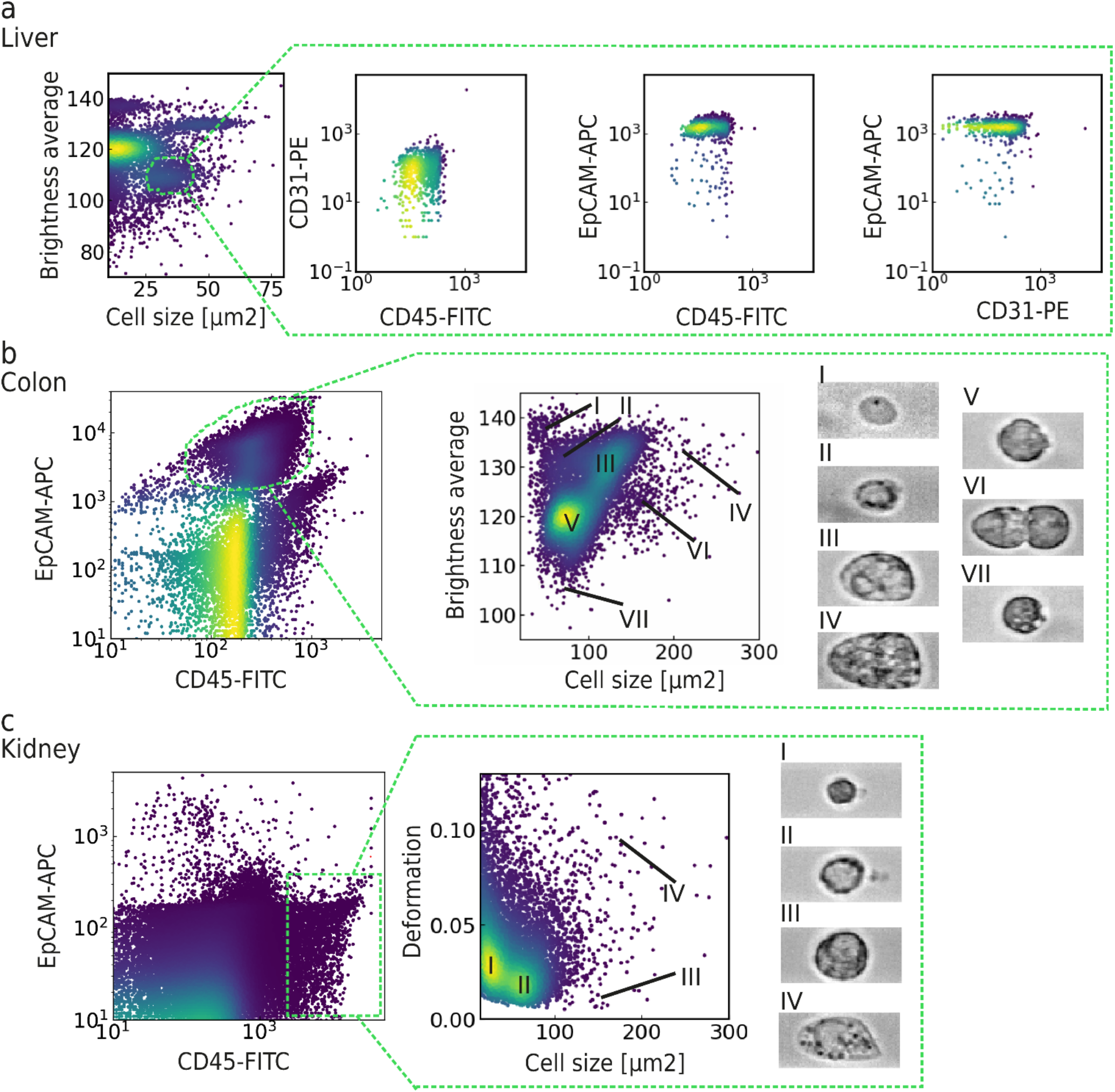
Representative scatter plots of physical parameters of cells from murine liver, colon and kidney samples. **a**, Representative scatter plots of brightness average vs cell size for cells isolated from the liver showing numerous clusters of cells. The marked population of cells (forming a distinct cluster at cell size 25-50 μm^2^ and brightness average 100-115) is enriched for EpCAM positive (epithelial) cells but devoid of endothelial cells or leukocytes. **b**, Representative scatter plots of colon cells stained for EpCAM and CD45 cell surface markers. Within the EpCAM positive population, 7 subpopulations of cells can be distinguished based on their physical parameters, *i*.*e*. brightness and cell size. **c**, Representative scatter plots of kidney cells stained for EpCAM and CD45. Within the CD45 population, 4 subpopulations can be distinguished based on cell size and deformation.

Fig. 2b and Fig. 2c highlight the advantage of using image-based physical phenotyping over relying on conventional fluorescence-based flow cytometry alone. In conventional flow cytometry it is hardly possible to distinguish individual subpopulations of epithelial (EpCAM+) cells unless extra panels of fluorescent antibodies against known and pre-defined cell types are used. Distinction of various subpopulations was possible with RT-FDC thanks to the extra information depth provided by physical phenotype parameters. Within the epithelial cells of colon, we identified 6 clusters of cells purely based on brightness and size (Fig. 2b). Similarly, within the leukocyte (CD45+) population of the kidney, we found 4 different clusters based on the cell size and deformation parameters (Fig. 2c). We note that, using the sorting modality recently developed for RT-FDC ^34^, any of these cell populations can be isolated according to the image-derived parameters and analysed for their molecular identity, *e*.*g*. by subsequent RNA sequencing.

RT-FDC can also be used to capture cell interactions. Using the aspect ratio and cell size parameters, we identified cell doublets in the thymus and spleen samples. Many doublets were composed of two different cell types, according to the cell surface markers (Supplementary Fig. 3). The position of the cell within the channel in combination with the position of the fluorescent peak allowed us to identify, for instance, that a doublet was composed of a leukocyte (CD45+) and an endothelial cell (CD31+) (Supplementary Fig. 3b and d). Using the RT-FDC sorting module ^34^, cell doublets can be isolated label-free for further molecular analysis and downstream applications, including studies of physically interacting immune cells in tissue ^35^.

We observed that RT-FDC analysis of single cell suspensions obtained from mechanical and enzymatic dissociation showed different population distributions for certain tissue types. For instance, the mechanical dissociation of liver and lung showed an enrichment of bigger cells (> 150 μm^2^ for liver, > 50 μm^2^ for lung) compared to enzymatically processed samples (Supplementary Fig. 4). In liver, cells ranging between 150 μm^2^ and 1000 μm^2^ in cross-sectional cell area (corresponding to 7-18 μm in radius) were determined as hepatocytes according to their morphology and size ^36^. As the major parenchymal cell type of the liver, hepatocytes account for 70% of the liver cell population and take up nearly 80% of liver volume ^37^. In the cell suspension obtained using TG, the proportion of hepatocytes to total cells was between 40-80%, showing that the yield of hepatocytes was nearly representative of their known presence in tissue. Mechanical dissociation seems less disruptive to sensitive cells such as hepatocytes which are extremely prone to cell death and are often lost during standard isolation procedures ^38^. Moreover, subpopulations of hepatocytes could be distinguished according to cell size (Supplementary Fig. 4b). We hypothesize that these populations correspond to hepatocytes of differing ploidy, as DNA content is strongly correlated with cell volume ^39^. Thus, our method could serve as a tool for the label-free monitoring of aging and pathophysiological processes in the liver, which are linked with the proportion of polyploid hepatocytes ^40^.

### Detecting the degree of tissue inflammation by the physical phenotyping of cells

Our method opened the possibility to investigate the changing physical phenotype of cells in solid tissues during disease progression. Previously, we studied physical phenotype changes of blood cells involved in inflammatory states ^22,25^ and thus decided to investigate the possibility to detect inflammation in solid tissue. Inflammatory bowel diseases (IBD), such as Crohn’s disease and ulcerative colitis, are chronic inflammatory disorders of the intestine associated with a compromised epithelial/mucosal barrier and activation/recruitment of immune cells ^41^. Although the aetiology of IBD is still not fully understood, much of our understanding about IBD comes from experimental animal models of intestinal inflammation. One such model is adoptive transfer of naïve T-cells into Rag1-deficient mice to induce experimental colitis (T cell transfer model of chronic colitis, from here on referred to as transfer colitis). The severity is then commonly quantified via a histopathological score generated from haematoxylin and eosin (H&E) stained slides of the colon tissue.

Our goal was to investigate changes in the physical phenotype of colon cells during transfer colitis. Scatter plots of deformation vs cell size suggest a difference between disease and healthy tissue (Fig. 3a), where cells from disease tissue appear less deformed than cells from healthy tissue. Upon examination of the CD45+ cells (Fig. 3b, c) it became evident that the transfer colitis samples were characterised by a high abundance of leukocytes with low deformation, likely lymphocytes. Overall, we found a significant decrease of median deformation in the transfer colitis samples, accompanied by a significant increase in the number of leukocytes, in accordance with infiltration of adoptively transferred lymphocytes (n = 14; Fig. 3d). The median deformation of cells was strongly negatively correlated with the number of leukocytes, with a Pearson’s correlation coefficient of *r(12)* = −0.69 (*p* = 0.0065; Fig. 3e). Furthermore, the median values of cell size and deformation were linked with expert H&E scoring (Supplementary Fig. 5); although correlation via linear fitting was not possible. Transfer colitis samples with high H&E score exhibited bigger cell size and lower deformation compared to healthy tissue.

**Fig. 3:**
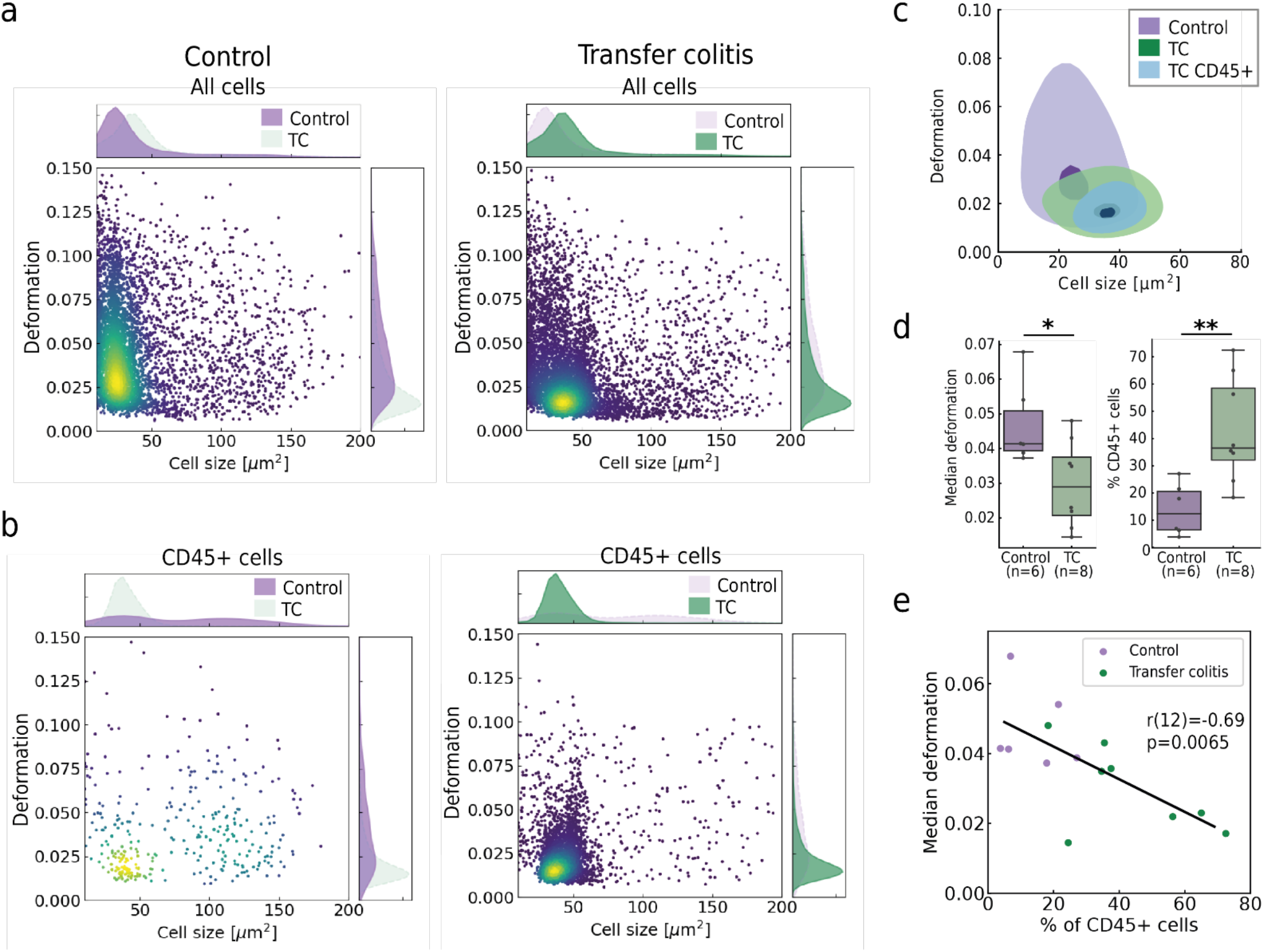
Physical phenotyping of cells via RT-FDC reflects tissue inflammation. **a**, Cell size vs deformation scatter plots of cells isolated from transfer colitis tissue samples (TC) compared to healthy murine colon tissue (Control); with corresponding cell size and deformation histograms. **b**, The same two colon samples gated for CD45 positive cells, showing the enrichment of leukocytes in transfer colitis samples, accompanied by changes of the physical phenotype parameters. **c**, Contour plots of samples shown in A and B. **d**, Quantification of median deformation and percentage of CD45 positive cells (n = 14). Statistical comparisons were performed using Student’s t-test, *p-*values are represented by * *p* < 0.05, ** *p* < 0.01, *** *p* < 0.001. **e**, Pearson’s correlation of median deformation and the proportion of leukocytes (CD45+ cells).

### The physical phenotype of cells is distinct in tumour versus healthy mouse colon tissue

Previous studies have found differences between the mechanical properties of cancer cells and their healthy counterparts ^11,12,42–45^. A major drawback of these studies is laborious sample preparation and low throughput that limits the conversion of these studies to actual diagnostic approaches. Given the rapidity of our approach to obtain and assay the mechanical phenotype of single cells from solid tissues, we explored its potential to detect colorectal cancer. We used mice deficient in an intestinal epithelial cell-specific protein with a key role in epithelial integrity. These animals spontaneously develop colon tumours. We examined a total of 17 mice and compared cells isolated from tumours to cells from a healthy part from the colon of the same animal. It is noteworthy that the healthy tissue was always more difficult to mechanically break apart into single cells, and tumour tissue yielded more intact cells. We analysed cells greater than 60 μm^2^ (determined by cross-sectional area), as below this threshold the sample comprised mainly of immune cells (Supplementary Fig. 6) and small debris.

Our results showed that the physical phenotype of cells from tumour tissue significantly differed from the control samples. Representative plots from a single mouse in Fig. 4a-c demonstrate that cells from the tumour had larger cell size and higher deformation than their healthy counterpart. The analysis of all 34 samples revealed that cells from tumours had significantly higher mean cell size (Fig. 4d), deformation (Fig. 4e), aspect ratio (Fig. 4f) and area ratio (Fig. 4g). The tumour samples also exhibited greater heterogeneity, demonstrated by the broader distribution in Fig. 4c and significantly higher standard deviations of cell size, deformation and area ratio (Fig. 4d-g).

**Fig. 4:**
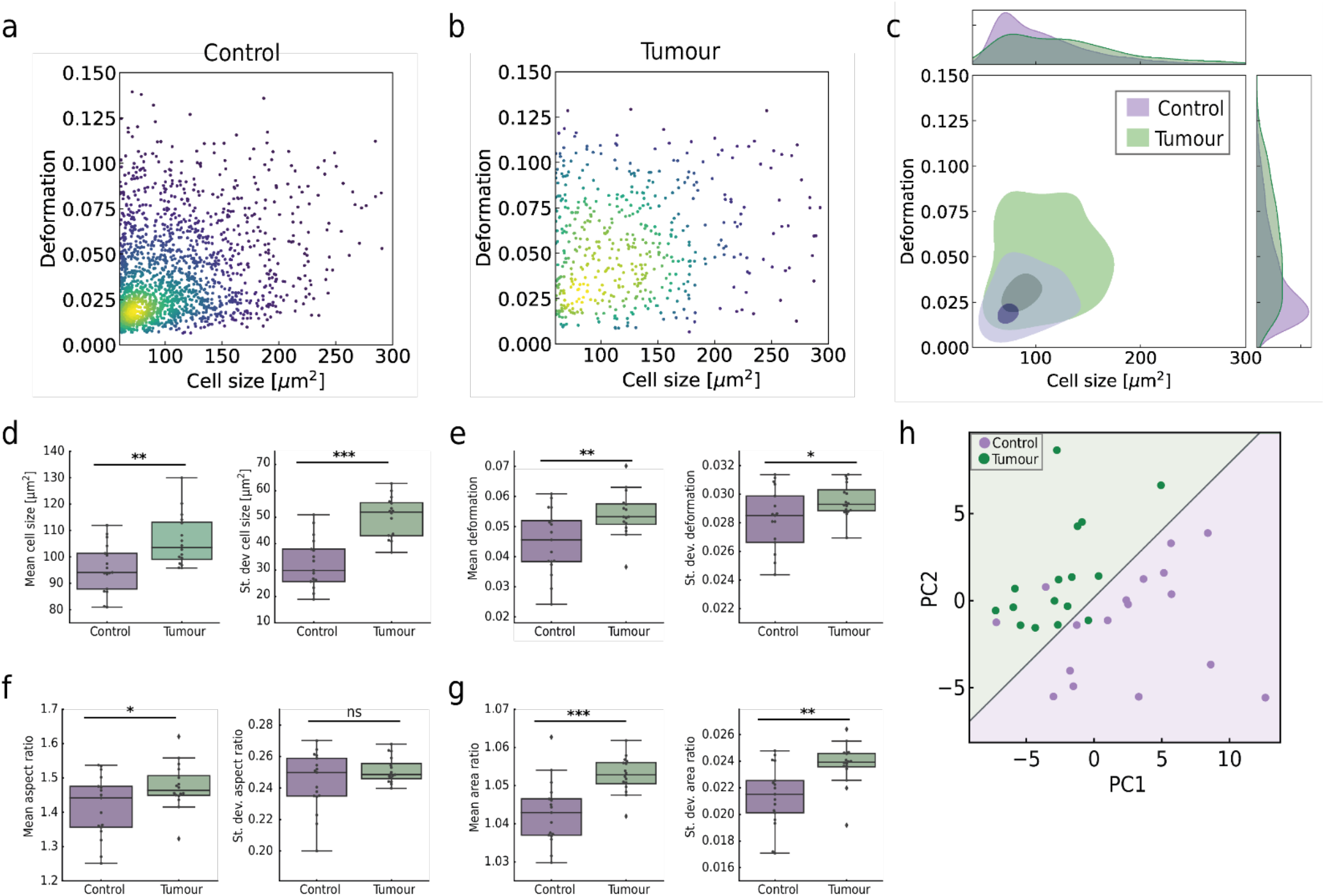
Physical phenotype of cells in tumorous and healthy mouse colon tissue. **a**, Cell size vs deformation scatter plots of a control sample of murine colon tissue compared to **b**, tumour tissue. **c**, The contour plot of samples shown in A and B, with corresponding cell size and deformation histograms demonstrating greater heterogeneity of cell size and deformation in tumour compared to the control tissue. **d-g**, Means and standard deviations of physical phenotype parameters of 17 control (purple) and 17 tumour samples (green); **d**, cell size, **e**, deformation, **f**, aspect ratio, **g**, area ratio. Statistical comparisons were performed using Student’s t-test, *p-*values are represented by * *p* < 0.05, ** *p* < 0.01, *** *p* < 0.001, ns = non-significant. **h**, Principal component analysis of mouse colon tissue samples, where green points represent tumour samples and purple points represent the control samples. Linear regression analysis was performed on PC1 and PC2 with the resulting two categories shown as purple (control) and green (tumour) background colours.

We next investigated whether the physical phenotype differences could be exploited for the reliable distinction between tumorous and healthy tissue. For this, we divided cells into three categories according to cell size (60-90 μm^2^, 80-120 μm^2^, 120-200 μm^2^). For each size category twelve parameters were derived: mean, median and standard deviations of the cell size, deformation, aspect ratio and area ratio; adding up to a total of 36 parameters for each sample (Supplementary Fig. 7). These parameters were subjected to principal component analysis (PCA), Fig. 4h. 58.5% of the variance was explained by the two principal components PC1 (39.8%) and PC2 (18.7%). The relative importance of physical features in determining the principal components is shown in Supplementary. Fig. 7. The most dominant feature for PC1 was the deformation of cells between 60 and 120 μm^2^, whilst in the case of PC2 cell size parameters prevailed. Logistic regression performed on the PCA (shown by the linear divide in Fig. 4h) demonstrated that the condensed physical phenotype information represented by the principal components suffices to distinguish between healthy and tumour tissue; 31 out of 34 samples lay in the correct region.

### Reliable distinction of tumour and healthy tissue in frozen or fresh human biopsies

We next sought to challenge our method for detecting tumours from human biopsy samples. As a first step, we performed RT-FDC analysis on cells isolated from 13 cryopreserved biopsy samples of colorectal cancer and 13 samples of healthy surrounding tissue from the same patients. PCA was performed on 45 parameters (Fig. 5a; Supplementary Fig. 8) with 41.7% of the variance explained by the two principal components (25.3% and 16.4% for PC1 and PC2, respectively). The PCA showed that tumour and healthy samples segregate along the PC2 mainly by the deformation and standard deviation of brightness of cells larger than 100 μm^2^ (Fig. 5a). Cell size parameters of cells under 100 μm^2^ also contributed to the separation of the samples. Of note, the selection of RT-FDC parameters had to be optimised to get a good separation between the healthy and tumour tissue. Logistic regression was performed on the PCA (shown by the linear divide in the PCA plot) and used to predict the classification of six blind samples (shown as crosses in Fig. 5a); all six samples were correctly classified as either healthy or tumour tissue. We examined the minimal number of cells needed for correct classification of these blind samples (Supplementary Fig. 9). Approximately 1,500 cells had to be analysed in a sample for correct classification, corresponding to a minimal RT-FDC measurement time of approximately 5 minutes.

**Fig. 5:**
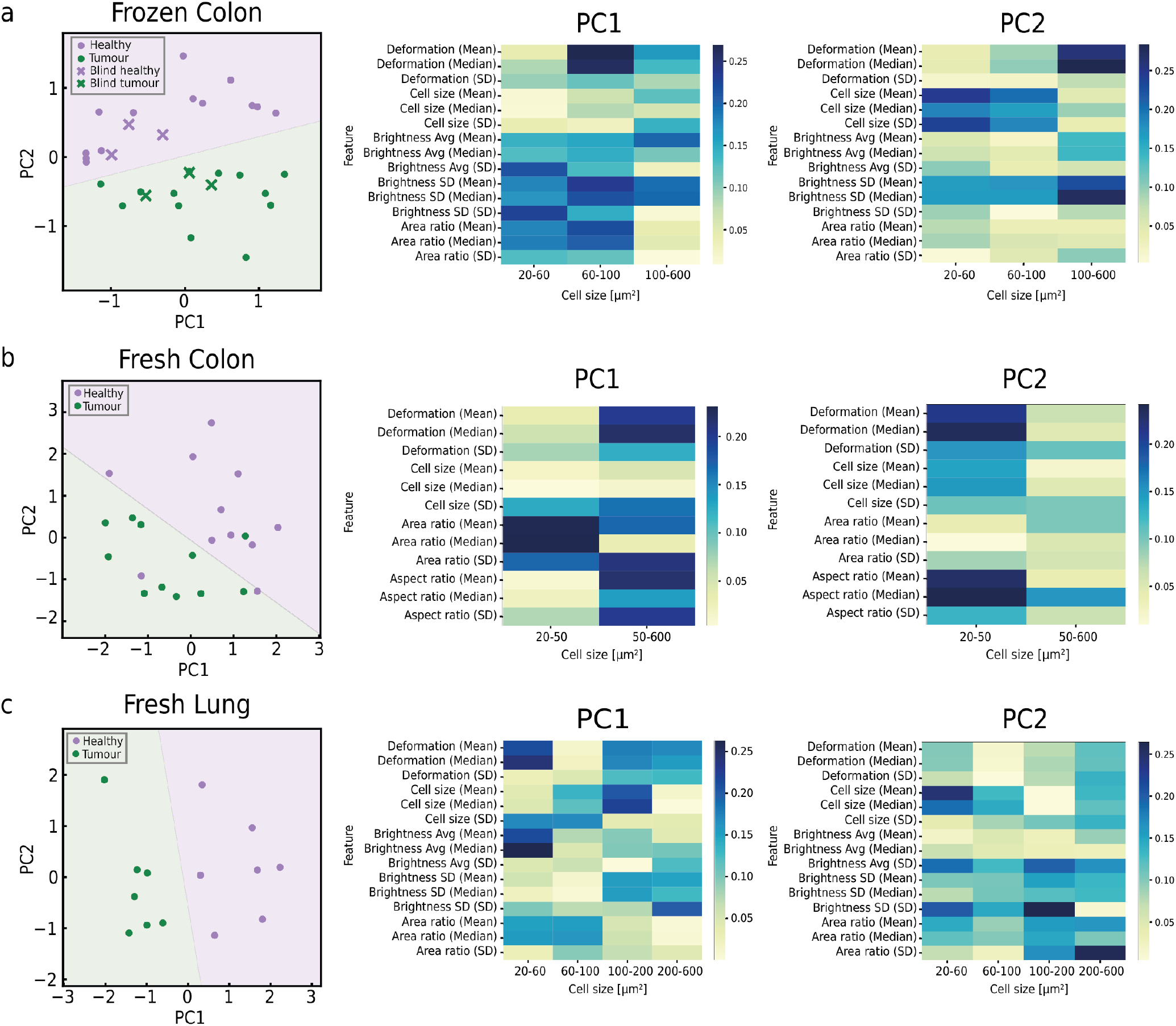
Distinction of tumour and healthy tissues in human biopsies using PCA and logistic regression. In the PCA plots on the left, green points represent tumour samples and purple points represent the healthy surrounding tissue. Logistic regression was performed on each of the PCAs with the resulting two categories shown as purple (healthy) and green (tumour) background colours. The feature importance analysis to the right of the PCA plot shows the colour-coded significance of each feature for determining PC1 and PC2 for that particular tissue; the x axis lists cell size categories; the y axis lists RT-FDC parameters and their statistical features derived across all samples (in brackets). **a**, PCA on RT-FDC parameters of 30 frozen colon samples (15 tumour biopsies and 15 samples of surrounding tissue). **b**, PCA on RT-FDC parameters of 22 fresh colon biopsy samples (11 tumour, 11 healthy). **c**, PCA on RT-FDC parameters of 14 fresh lung biopsy samples (7 tumour, 7 healthy).

The short-combined tissue processing and analysis time (< 30 minutes) opened up the applicability of the method for intraoperative pathology, the examination a patient’s biopsy sample during surgery. To explore this idea, we analysed freshly excised biopsies samples from colorectal cancer patients (n = 11). The algorithm trained on frozen colon tissue did not perform well for fresh tissue, possibly due to differences of physical phenotype between the frozen and fresh tissue (Supplementary Fig. 10), as well as the absence of certain cell types lost during the freezing process. Therefore, new PCA was performed on data from fresh colon biopsies in two size categories (20-50 μm^2^ and 50-600 μm^2^) (Fig. 5b), where 68.5% of the variance was explained by the two principal components (36.5% and 32% for PC1 and PC2, respectively). Here, the deformation of cells contributed strongly in the PCA and cell size was less important than the other physical phenotype parameters. Upon logistic regression, 3/22 samples were not correctly classified, which could be attributed to inter-tumour or intra-tumour heterogeneity. Nevertheless, using our approach we achieved 100% accuracy in classifying healthy and tumour samples from frozen biopsies, and 86% accuracy for the separation of fresh biopsy samples.

To validate our method on tissue from a different organ, we applied it to freshly excised lung biopsy samples from seven cancer patients. PCA combined with logistic regression easily separated all healthy from tumour samples (Fig. 5c). In the PCA, 46.9% of the variance was explained by PC1 and PC2 (31.2% and 15.7%, respectively). Deformation parameters contributed strongly to PC1, which again demonstrates that the unique information brought by cell deformability measurement is useful for distinguishing between tumorous and healthy tissue, and the technique should be considered and adapted for routine clinical practice.

## Discussion

Here, we present a quick and simple method for the processing and analysis of cells from solid tissue, suitable for biopsy diagnostics. To our knowledge, this is the first demonstration of the diagnostic potential of physical phenotyping of single cells from solid tissue samples which is applicable in practice. Mechanical dissociation of tissue within a few minutes is followed by high-throughput analysis of cells in deformational flow. Within a few minutes, thousands of cells are imaged and various physical phenotype features are extracted from each cell image. The method is label-free and relies simply on brightfield images, in contrast to molecular diagnostic tools or conventional flow cytometry, where expensive reagents or fluorescent markers are needed. Importantly, the information is available within 30 minutes of biopsy excision, which can be an advantage when there is necessity to detect pathology quickly. This is the case during intraoperative consultation that provides diagnostic information during cancer surgery and often defines the further course of the procedure. The standard workflow requires transportation of the biopsy sample to the pathology department, where it is embedded in a mounting medium (optimal cutting temperature compound), frozen and cut in thin slices using a cryostat. The slides are then prepared with H&E staining, and pathologists assess numerous characteristics including the nature of the lesion (*i*.*e*. its malignancy) ^29,46^. Our workflow circumvents the freezing and staining steps, could be performed directly on site and allows to detect malignancy based on the automated assessment of physical parameters of single cells.

Beyond intraoperative diagnosis, we show that our method is useful for rapid examination of IBD samples. Clinical diagnosis of IBD in most cases requires the combination of different tests including a blood test, stool examination, endoscopy and histological analysis of mucosal biopsies ^47,48^. Histological scoring has growing importance in the IBD field, as the histological level of inflammation correlates with recurrence of disease, probability of surgery and risk of cancer. Our study shows that the degree of tissue inflammation in a colon biopsy sample can be obtained by monitoring cell physical phenotype via RT-FDC, bypassing the need for staining or expert’s assessment. We envision that the method could be used to monitor temporal inflammatory changes to assess disease progression and response to treatment, and to provide an objective diagnostic scoring system for daily clinical practice, which is currently lacking in the IBD field.

Previous studies on cancer cells have shown a strong correlation between malignancy and the mechanical properties of cells ^10–12,42,45,49^. Here, we exploit this correlation for detecting malignancy in human tissue biopsies. RT-FDC probes cell deformability, at a high-throughput rate, by exposing cells to shear flow in a microfluidic channel and allows the mechanical phenotyping of single cells, using an analytical model and numerical simulations ^50,51^. Assuming an initial spherical cell under normal (stress-free) conditions, RT-FDC can provide a quantitative measure of an elastic modulus. However, in heterogenous tissue samples, such as the ones used in this study, the cells are often not spherical prior to entering the microfluidic channel. Nevertheless, the degree of deformation in this standardized deformation-assay can be interpreted as a qualitative measure of deformability and the deformation information inherent in the images is shown to be valuable diagnostic marker. PCA of murine colon samples and human colorectal biopsies revealed that cell deformation in standardised channel flow conditions is key for distinguishing between healthy and tumorous tissue. This highlights the uniqueness of the information brought by this method, currently missing from routine diagnostic practices which to date rely mostly on histological assessment.

For practical clinical use, it will be necessary to integrate the tissue processing and single-cell phenotype analysis into a single automated pipeline. Although mechanical dissociation using a tissue grinder is an efficient way to obtain single cells from tissues for diagnostic application, it will be important to reduce the manual handling steps such as filtering and concentrating cells. However, even in its current state, it is faster and more cost-effective than enzymatic processing of tissue. A key advantage of mechanical dissociation was that the processing time took less than 5 minutes per sample, as opposed to tens of minutes or even several hours for enzymatic dissociation protocols. Moreover, enzymatic protocols typically require sample-dependent reagents that are often expensive and require special storage conditions, while mechanical dissociation can be performed in standard culture medium. Whereas different enzymatic protocols often enrich for specific cell types ^52^, we believe that the single-cell suspension from mechanical dissociation might be more representative of the actual populations in tissue and is therefore suitable for an unbiased examination of the cellular landscape. Fast dissociation also has the potential to preserve biochemical and biophysical properties of cells in a state near to *in situ*, properties which likely deteriorate with increased processing times in other approaches. Due to the speed of the mechanical dissociation, cells might undergo less proteomic or transcriptional changes, which are known to happen during enzymatic processing ^32,53–56^. Further comparative and molecular studies are necessary to assess these assumptions.

In the future, investigation on larger patient cohorts will allow to exploit machine learning for diagnostic or prognostic decision making. Artificial intelligence (AI) is a powerful tool for diagnostic tasks, aiding pathologists in inspecting histological whole slide images, diagnosing cancer or classifying tumours ^57–59^. The large datasets obtained by RT-FDC analysis, composed of thousands of cell images and multidimensional information, lend themselves for such AI approaches. In this study, we focused on parameters calculated from images in real-time, but additional physical phenotype parameters can be calculated post acquisition and used as further inputs for machine learning, *e*.*g*. shape or texture features. Future work will also focus on the correlation between the physical phenotype data and tumour malignancy scoring, metastatic potential and survival rate.

Finally, an important aspect of the method is that the physical phenotype of cells can be used to identify cell populations in tissue in a label-free manner. This renders our approach unbiased compared to conventional fluorescence-based analysis, which relies on prior knowledge of the molecular signature of a specific population within a heterogenous mixture of cells. In simple terms, one does not need to know which cell types to expect prior to analysis. Furthermore, thanks to the sorting modality recently added to RT-FDC ^34^, a specific population of cells can be isolated according to parameters calculated from images in real-time or using trained neural networks. In the future, this could be employed for enrichment of uncharacterised cell populations in tissue for downstream OMICS analysis or even for regenerative medicine purposes, *e*.*g*. for the label-free isolation of tissue-derived stem cells.

Taken together, our findings demonstrate the potential of physical phenotyping of cells via RT-FDC after enzyme-free mechanical tissue dissociation as a quick and simple method for diagnosing pathological states in tissue biopsies, in particular to provide a rapid and unbiased prediction of disease state in inflammatory conditions and in malignancy.

## Methods

### Animal experiments

All animal experiments were conducted in collaboration with the Department of Internal Medicine 1, University Hospital Erlangen, in compliance with all institutional guidelines. Animal studies were conducted in a gender and age-matched manner using littermates for each experiment. Both male and female animals were used. All mice were kept under specific pathogen–free conditions. Mice were routinely screened for pathogens according to FELASA guidelines (TVA No: 55.2.2-2532-2-1032/ 55.2.2-2532-2-473). Animals were euthanised by cervical dislocation and organs were surgically removed. Lung and liver tissues perfusion preceded the enzymatically dissociation process. For mechanical dissociation using a tissue grinder, organs were washed thoroughly with PBS before being placed in DMEM supplemented with 2% FBS and placed on ice until further processing. Enzymatical protocols were obtained from literature and are summarised in Supplementary Table 1.

### Spontaneous tumour model

To generate a specific deletion of an intestinal epithelial cells-specific protein with a key role for epithelial integrity, mice carrying LoxP-Cre flanked for the specific protein were crossbred with VillinCre mice. Spontaneous tumorigenesis was observed in colon with 100% of penetrance.

### Adoptive lymphocyte transfer colitis

Immunodeficient Rag1-/- mice received 1 million of CD4+CD25- T cells via i.p route. Mononuclear cells were isolated from the spleen of C57/BL6 donor mice and purified using MACS technology, before being injected into immunodeficient mice. Animals were sacrificed three weeks upon cell transfer and the colon tissue was processed as described in the Tissue dissociation and single cell preparation section.

### Human tissue preparation

Surgically resected human biopsy samples (obtained from the Pathology Institute, Erlangen) from tumour or healthy tissue were immediately placed in DMEM Advanced medium supplemented with 10% Foetal Bovine Serum (FBS), 1% GlutMAX™,1% HEPES and 1% penicillin/streptomycin and stored at 4°C, processed immediately or frozen in liquid nitrogen for later use. This study is covered by ethic votes of the University Hospital of the Friedrich-Alexander University Erlangen-Nürnberg (24.01.2005, 18.01.2012).

### Tissue dissociation and single cell preparation

Tissue dissociation using a TissueGrinder (TG; Fast Forward Discoveries GmbH) was performed, as described in ^32,33^. Briefly, the tissue sample was cut into small pieces of about 1-2 mm and placed into the rotor unit of the TG with 800 μl of DMEM supplemented with 2% FBS. The rotor unit was positioned in the lid of a 50 ml Falcon tube; the stator insert with a 100 μm cell strainer was placed on top of the rotor unit. A 50 ml falcon tube was placed on the lid, screwed and positioned on the TissueGrinder device (Fig. 1). The grinding process parameters for each tissue type are summarised in Supplementary Table 2. TG protocols were provided by the manufacturer with some minor modifications ^32,33^. Following the grinding procedure, the Falcon tube was inverted onto a rack, opened and the cell strainer washed with 5 ml of DMEM, 2% FBS. The flow through was transferred into a 15 ml Falcon tube and centrifuged for 8 minutes at 300 *x g*. Subsequently the supernatant was aspirated, and the cell pellet washed with 2 ml of PBS, 2% FBS, passed through a FACS tube with a cell strainer cap and centrifuge at 300 *x g* for 5 minutes. The cell pellet was resuspended in a high viscosity measurement buffer prepared using 0.6% (w/w) methyl cellulose (4,000 cPs; Alfa Aesar) diluted in phosphate buffer solution (PBS) without calcium and magnesium, adjusted to an osmolality of 270-290 mOsm/kg and pH 7.4. The viscosity of the buffer was adjusted to (25±0.5) mPa·s at 24 °C using a falling sphere viscometer (HAAKE Falling Ball Viscometer Type C, Thermo Fisher Scientific).

### Real-time fluorescence and deformability cytometry

RT-FDC measurements were performed as previously described ^13,14^, using an AcCellerator instrument (Zellmechanik Dresden GmbH). The cell suspension was drawn into a 1 ml Luer-Lok syringe (BD Biosciences) attached to a syringe pump and connected by PEEK-tubing (IDEX Health & Science LLC) to a microfluidic chip made of PDMS bonded on cover glass. A second syringe filled with pure measurement buffer was attached to the chip and used to hydrodynamically focus the cells inside the constriction channel. The microfluidic chip consists of a sample inlet, a sheath inlet and an outlet connected by a central channel constriction of a 20×20 μm, 30×30 μm or 40×40 μm square cross-section and a length of 300 μm. The corresponding total flow rate used: 0.06 μl/s, for 20 μm channel; 0.12 μl/s, for 30 μm channel; 0.2 μl/s, for 40 μm channel; with a sheath to sample flow ratio of 3:1. The chip was mounted on the stage of an inverted high-speed microscope equipped with a CMOS camera. The laser power for each fluorophore was adjusted accordingly, based on single stain controls and an unstained sample. An image of every cell was captured in a region of interest of 250 × 80 pixels at a frame rate of 2000 fps. Morphological, mechanical and fluorescence parameters were acquired in real-time. The fluorescence threshold for each antibody was adjusted according to an unstained sample of cells obtained from the same tissue. Supplementary Table 3 lists the features acquired in real-time and during post processing analysis; described in detail in previous publications ^60,61^.

The calculation of deformation, a measure of how much the cell shape deviates from circularity, was obtained from the image using the projected area (*A*) and cell contour length calculated from the convex hull (*l*):

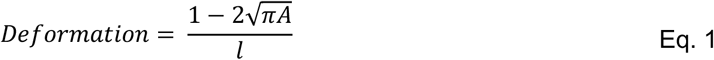

### Fluorescence labelling

Where necessary, single cell suspensions were incubated for 20 minutes at room temperature with 200 μl of corresponding antibodies (Supplementary Table 4 for antibodies dilution) diluted in PBS supplemented with 0.5% Bovine Serum Albumin (BSA, Sigma-Aldrich) and FcR Blocking reagent of corresponding species (Miltenyi Biotec, Human: 130-059-901; Mouse: 130-092-575). The antibodies were washed by adding 1 ml of PBS, 2% FBS and centrifuged for 500 x g for 5 minutes. The final cell preparation was then resuspended in the measurement buffer before loading onto the microfluidic chip for RT-FDC analysis. For frozen biopsy samples, the tissue was placed in pre-warmed DMEM, supplemented with 10% FBS for 10 minutes and allowed to thaw prior to processing as described above.

### Data analysis

Cell images were analysed using Python 3.7. Dclab library was used for the initial loading, pre-processing and filtering of RT-FDC data ^62^. In order to remove images of debris, damaged cells and red blood cells, we applied gates for minimum cross-sectional area (20 μm^2^), area ratio (1-1.1) and aspect ratio (1-2).

Scikit learn package was used for further data processing and analysis ^63^. Parameters obtained from RT-FDC were transformed by scaling each feature to the range between (0,1). PCA was used for linear dimensionality reduction, using Singular Value Decomposition of the data to project onto a two-dimensional space (PC1 vs PC2). Logistic Regression was used for the classification of healthy vs tumour samples.

Statistical analysis was done in Python 3.7. Paired Student’s *t*-test was used to assess differences between fresh colon tissue samples and the corresponding frozen samples. The rest of statistical analyses were performed using independent Student’s t-tests. In graphs, *p-*values are represented by * *p* < 0.05, ** *p* < 0.01, *** *p* < 0.001. Pearson’s correlation was performed to judge the correlation between cell deformation and the number of CD45+ cells.

## Data Availability

The RT-FDC datasets generated and analysed for Fig 2, 3 and Supplementary Fig. 2 and 3 are available on figshare [https://figshare.com/s/2b3a1e1441f59813c24f]. Due to the size restrictions on the figshare repository, the RT-FDC datasets generated from human biopsies samples and mouse tumour samples are available from the corresponding author on reasonable request.

https://figshare.com/s/2b3a1e1441f59813c24f

## Data availability statement

The RT-FDC datasets generated and analysed for Fig 2, 3 and Supplementary Fig. 2 and 3 are available on figshare [https://figshare.com/s/2b3a1e1441f59813c24f].

Due to the size restrictions on figshare repository, the RT-FDC datasets generated from human biopsies samples and mouse tumour samples are available from the corresponding author on reasonable request.

## Code availability statement

The Python code for analysing the RT-DC datasets and performing the PCA analysis is available from the corresponding author on reasonable request.

## Acknowledgments

We thank R. Grützmann and K. Bende (Universitätsklinikum Erlangen) for kindly assisting us with biopsy samples. We also thank J. Kayser and M. Urbanska for critical revision of the manuscript. We acknowledge financial support from the DFG (SFB/TRR241 „IBDome“ to R.LP., M.W., R.A. and M.N.; FOR2438 to M.N; DI 1537/20-1, DI 1537/22-1, SFB CRC1181 and SFB TR221 to J.D.) and the IZKF Erlangen (research grant A79 to J.D.).

## Author contributions

M.K., D.S. and J.G. conceived the study and managed the project progress. D.S. and M.K. coordinated the experiments and analysis. D.S. developed experimental protocols and performed experiments together with C.S.. Data analysis and visualisation was performed by M.K. Other authors assisted with the experiments and participated in critical discussions. M.K. and D.S. wrote the manuscript with all authors providing feedback.

---

**Supplementary Table 1:** Enzymatic or mechanical dissociation protocols used for each organ

| **Tissue** | **Enzymatic/Mechanical Dissociation Procedure** |
| --- | --- |
| **Colon** | Miltenyi Biotec Lamina Propria Dissociation Kit |
| **Small Intestine** | Miltenyi Biotec Lamina Propria Dissociation Kit |
| **Lung** | Miltenyi Biotec Lung Dissociation kit |
| **Liver** | Miltenyi Biotec Liver Dissociation Kit or Collagenase/Dispase enzymatic cocktail |
| **Stomach** | Ruiz et al 2012 (Ruiz et al., 2012) |
| **Kidney** | Valente et al 2011 (Valente et al., 2011) |
| **Pancreas** | Epshtein et al 2017 (Epshtein et al., 2017) |
| **Thymus** | Mash tissue between frosted ends of two microscope slides |
| **Mesenteric Lymph Node** | Mash tissue between frosted ends of two microscope slides |
| **Spleen** | Mash tissue between frosted ends of two microscope slides |

**Supplementary Table 2:** Mouse organs processed with corresponding TG protocol used for mechanical dissociation and microfluidic chip size used for RT-FDC measurements.

| Tissue | TG protocol | Microfluidic chip size |
| --- | --- | --- |
| Colon | Colon | 20 µm |
| Small Intestine | Intestine | 20 µm |
| Lung | Lung standard | 30 µm |
| Liver | Liver standard | 30 µm & 40 µm |
| Kidney | Medium | 30 µm |
| Pancreas | Soft | 30 µm |
| Thymus | Thymus Soft | 30 µm |
| Mesenteric Lymph Node | Lymph nodes standard | 30 µm |
| Spleen | Thymus Soft | 30 µm |
| Human tumour | Human Tumour | 20 µm |

**Supplementary Table 3:** Parameters derived from cell images in real-time during an RT-FDC measurement.

| RT-FDC Parameters | Description |
| --- | --- |
| Area (cell size; $\mu\text{m}^2$) | The projected cross-sectional area of the cells is obtained from the number of pixels within the boundary of the cell, defined by the contour |
| Bounding box size x and y | A bounding box around the whole cell defines its length in x and y direction |
| Aspect ratio | Ratio between object's length (x) and height (y) |
| Circularity | Parameter that relates area (A) to the perimeter (P) of an object based on<br>$C = \frac{2\sqrt{\pi A}}{P}$ |
| Deformation | $\frac{1 - 2\sqrt{\pi A}}{l}$ |
| Inertia ratio | Ratio of the second moment of area over the y- and x-axis of the contour. |
| Area ratio | Ratio between the area of the convex hull and the area of the contour |
| Brightness [a.u.] | Mean of all pixel intensities inside the contour |
| Standard deviation of brightness | Standard deviation of the brightness of pixels inside the contour |
| X position | Position along the flow axis in the field of view. |
| Y position | Position perpendicular to flow axis in the field view |
| Volume | The apparent volume based on the assumption of rotational symmetry over the flow axis. |
| Elastic modulus (Young's Modulus) | Describes cell stiffness, based on assumptions of a model |
| Fluorescence area of peak [$\mu\text{s}$] | Area of the fluorescence intensity peak |
| Fluorescence maximum [a.u] | Maximum fluorescence intensity |
| Fluorescence width [$\mu\text{s}$] | Full width at half-maximum; used to identify the sub-cellular distribution of fluorescence marker |

**Supplementary Table 4:** Antibodies and fluorescent probe solutions used with the corresponding dilutions

|  | Reactivity | Dilution | Clone | Cat. No | Company |
| --- | --- | --- | --- | --- | --- |
| **CD326 (EpCAM) Alexa Fluor® 488** | Human | 1 in 100 | 9C4 | 324210 | BioLegend |
| **CD31(PECAM-1) APC** | Human | 5μl/reaction | WM59 | 303115 | BioLegend |
| **CD45 PE** | Human | 1 in 500 | HI30 | 304008 | BioLegend |
| **CD326 (EpCAM) FITC** | Human | 1 in 100 | 9C4 | 324203 | BioLegend |
| **CD45 Alexa Fluor® 700** | Human | 5μl/reaction | HI30 | 304024 | BioLegend |
| **CD45 Alexa Fluor® 700** | Mouse | 1 in 1000 | 30-F11 | 103128 | BioLegend |
| **CD326 (EpCAM) Alexa Fluor® 488** | Mouse | 1 in 200 | G8.8 | 118210 | BioLegend |
| **CD45 FITC** | Mouse | 1 in 150 | 30-F11 | 11-0451-82 | Thermo Fischer Scientific |
| **CD326 (EpCAM) APC** | Mouse | 1 in 250 | G8.8 | 17-5791-82 | Thermo Fischer Scientific |
| **CD31 (PECAM-1) PE** | Mouse | 1 in 1000 | 390 | 12-0311-82 | Thermo Fischer Scientific |
| **DRAQ5™** | - | 5μl/reaction | - | 62251 | Thermo Fischer Scientific |
| **Propidium Iodide** | - | 1 in 2000 | - | P1304MP | Thermo Fischer Scientific |
| **7-AAD Viability Solution** | - | 5μl/reaction | - | 420404 | Thermo Fischer Scientific |

