## Supplementary Figures for "Single-cell physical phenotyping of mechanically dissociated tissue biopsies for fast diagnostic assessment"

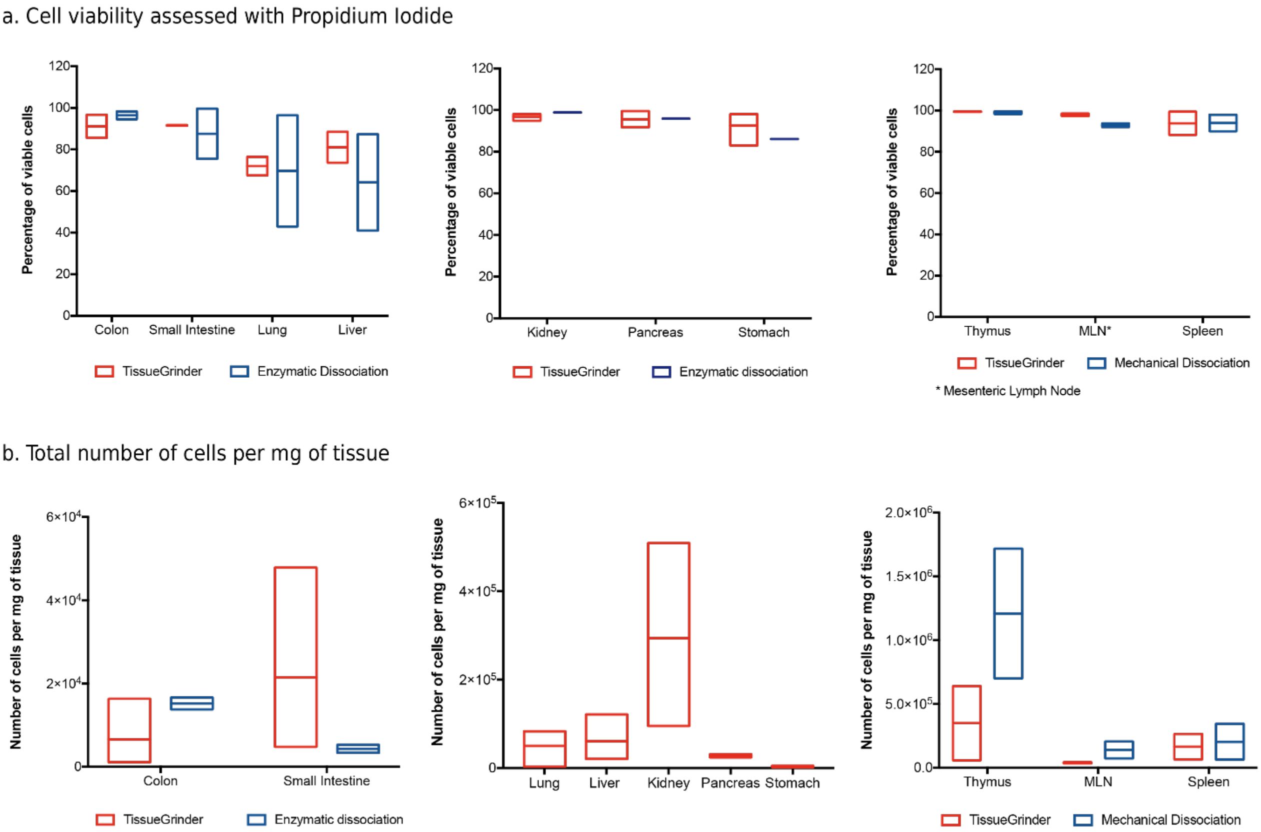


**Supplementary Fig. 1: Comparison of cell viability and cell yield of mechanical vs enzymatic dissociation of different murine tissues. a,** Percentage of viable cells for different organs dissociated using a tissue grinder (TG; marked in red) or enzymatic dissociation (marked in blue). Cell viability was assessed using propidium iodide and RT-FDC. **b,** Number of cells (obtained using a cell counter device) per mg of tissue processed. Lung, liver, kidney, pancreas and stomach processed with enzymatic dissociation were not weighted prior to the experiments. The line represents the mean ±SD (for TG data kidney and stomach: n = 3, pancreas n = 2; for enzymatic dissociation kidney, pancreas and stomach: n = 1; all other organs n = 2).

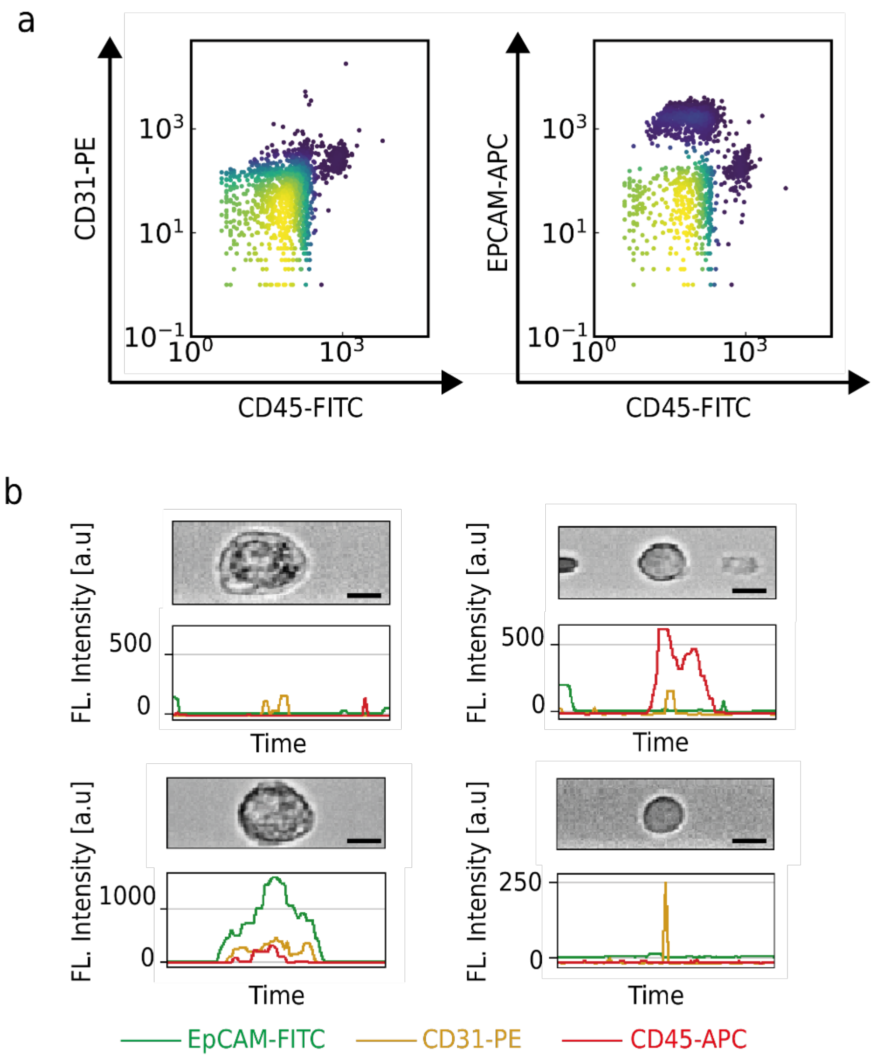


**Supplementary Fig. 2: Detection of fluorescent cell surface markers using RT-FDC. a,** Representative scatter plots of fluorescence intensities in three different detection channels of RT-FDC, showing the possibility of using fluorescent cell surface markers to characterise the cells. Plots show expression of an endothelial marker (CD31-PE) vs leukocyte marker (CD45-FITC); and an epithelial marker (EpCAM-APC) vs CD45-FITC. **b,** Representative images of cells and their corresponding fluorescent traces; the temporal shape of the fluorescence peak corresponds to the subcellular localization of the fluorophore. Top left to right: cell negative for all markers, a cell positive for CD45; bottom left to right: cell positive for EpCAM only and cell positive for CD31 only.

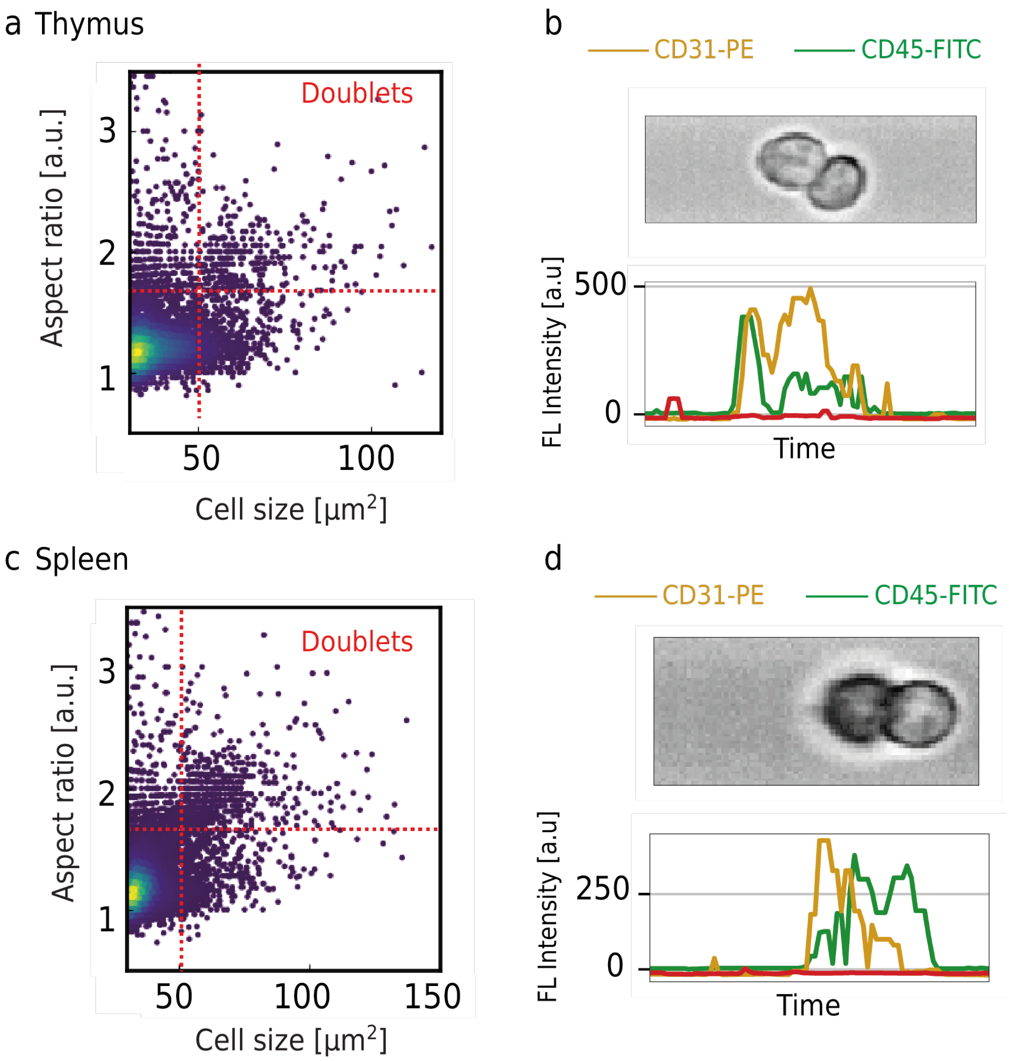


**Supplementary Fig. 3: Detection of cell doublets using RT-FDC.** Representative scatter plots of aspect ratio vs cell size of cells isolated from murine **a,** thymus and **c,** spleen showing the gating strategy for identifying cell doublets. Cell doublets identified in **b,** thymus and **d,** spleen with corresponding fluorescent traces, showing a leukocyte (CD45) attached to an endothelial cell (CD31).

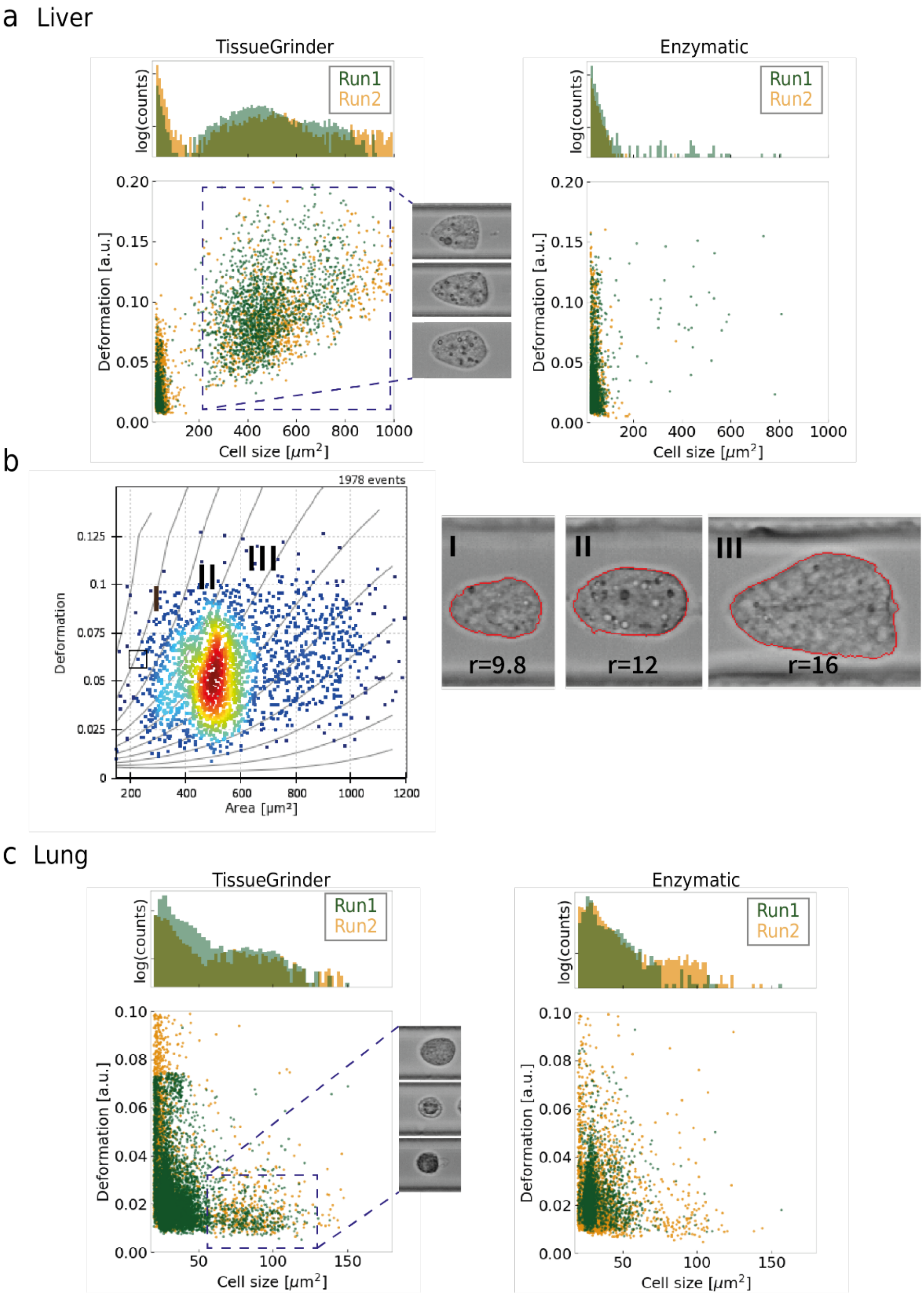


**Supplementary Fig. 4: Physical phenotype characterisation of cells isolated mechanically and enzymatically from murine liver and lung tissues. a,** Scatter plots of deformation vs cell size for cells isolated from mouse liver tissue using a tissue grinder or enzymatic dissociation. The selected cluster represents the enrichment of hepatocytes following mechanical dissociation, with representative images of the cells. **b,** Scatter plot of deformation vs cell size showing 3 clusters of cells that correspond to hepatocytes of different sizes; r = radius of cells**. c,** Scatter plots of deformation vs cell size for cells isolated from mouse lung tissue. The selected cluster represents the enrichment of bigger cells following mechanical dissociation, with representative images of the cells. Histograms represent the cell size distribution for each run. The green and orange colours represent two separate runs.

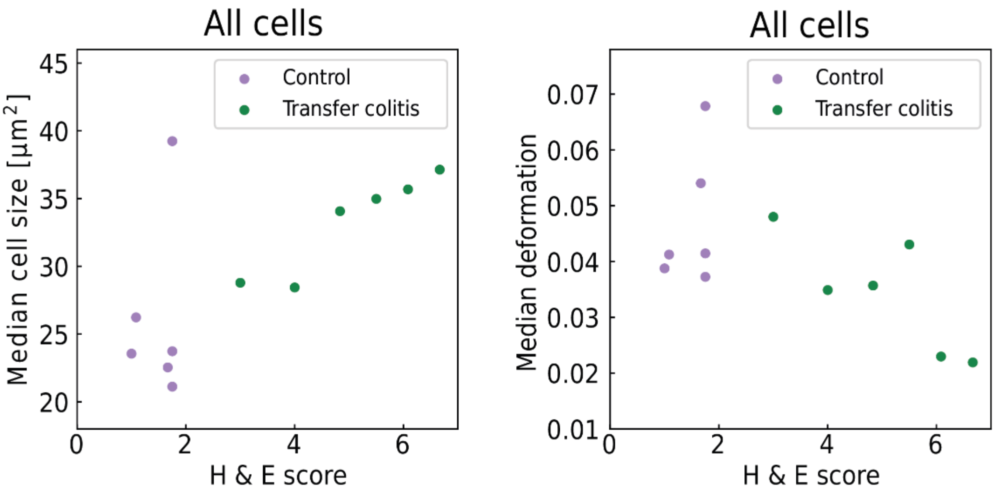


**Supplementary Fig. 5: Correlation of physical phenotype parameters obtained using RT-FDC and histopathology scoring of mouse transfer colitis samples.** Plot of Haematoxylin & Eosin (H&E) scoring vs **a,** median cell size and **b,** deformation of all cells measured. Transfer colitis samples are shown in green, control (healthy) samples are shown in purple.

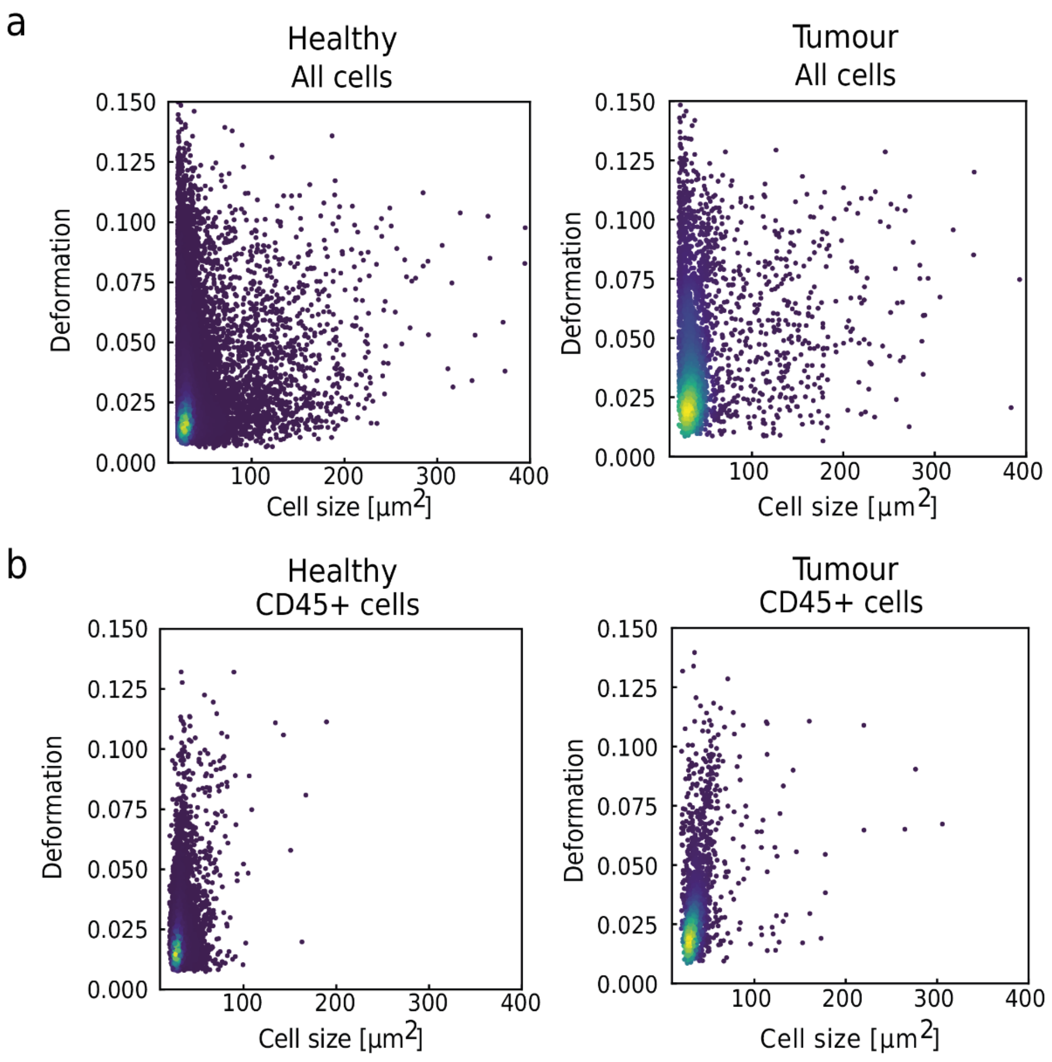


**Supplementary Fig. 6: Exclusion of small cells from analysis. a,** Representative RT-FDC scatter plots of deformation vs cell size, showing cells isolated from tumour or healthy murine colon samples. **b,** Representative scatter plots of CD45 positive cells (a marker of leukocytes), showing that a high percentage of these cells are less than 60 $\mu m^2$ in size. These cells were excluded from the analysis.

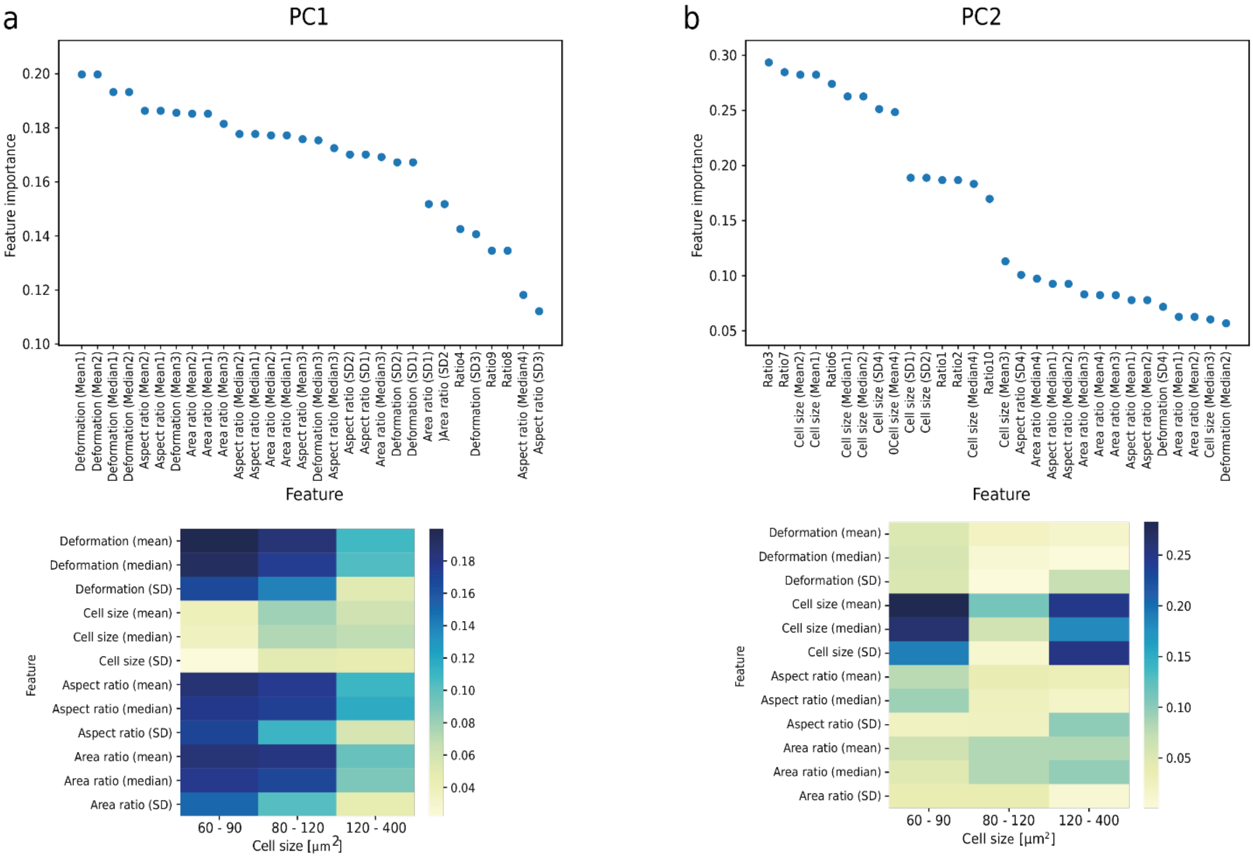


**Supplementary Fig. 7: Principal component feature importance for mouse tumours.** PCA was performed on a total of 36 parameters; 12 parameters (y axis) for 3 cell size categories (x axis). **a,** Relative feature importance for PC1 and **b,** PC2.

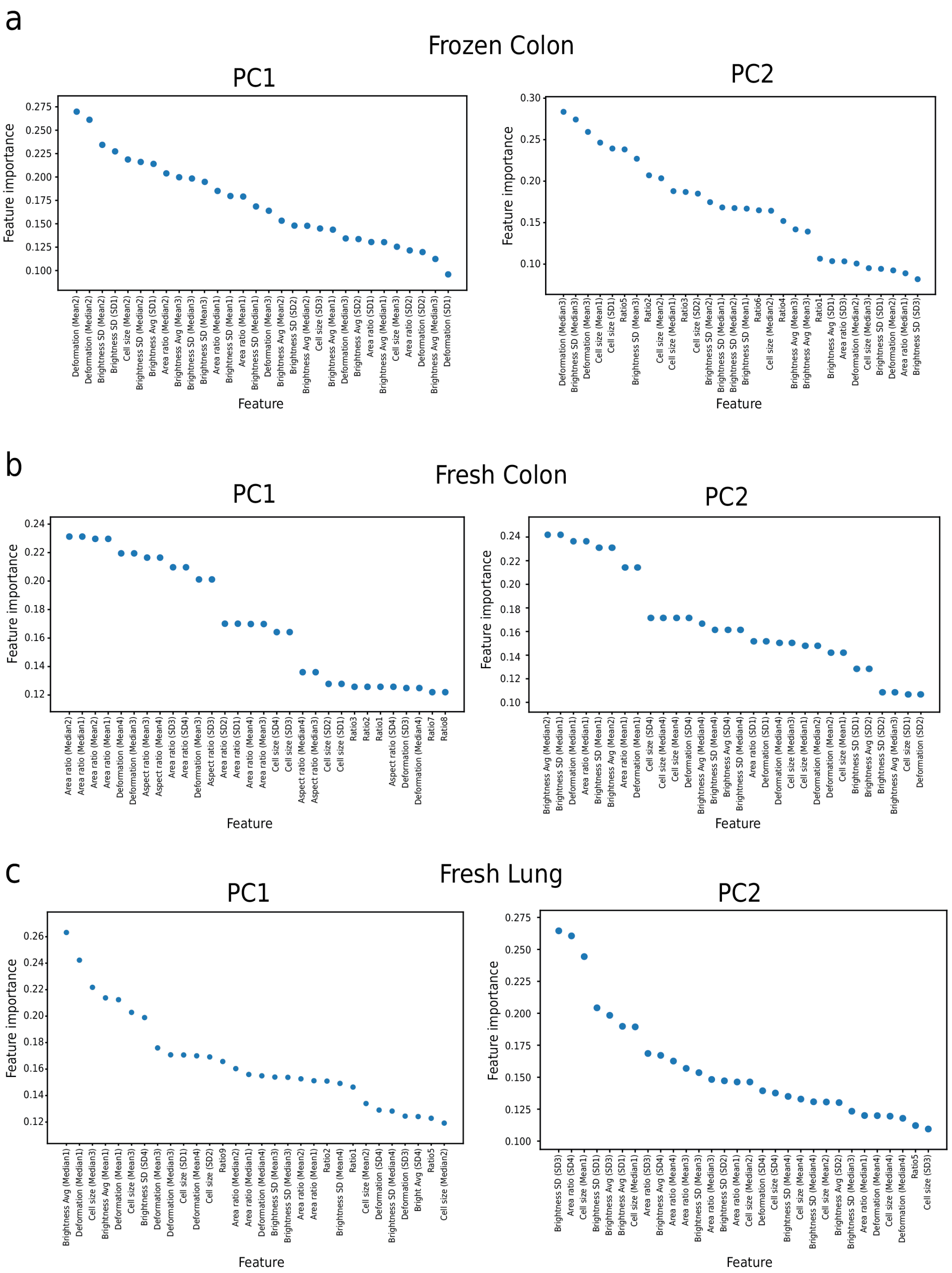


**Supplementary Fig. 8: Principal component feature importance for human biopsy samples.** Relative feature importance for PC1 and PC2 for **a,** frozen colon biopsy samples, **b,** fresh colon biopsy samples, **c,** fresh lung biopsy samples.

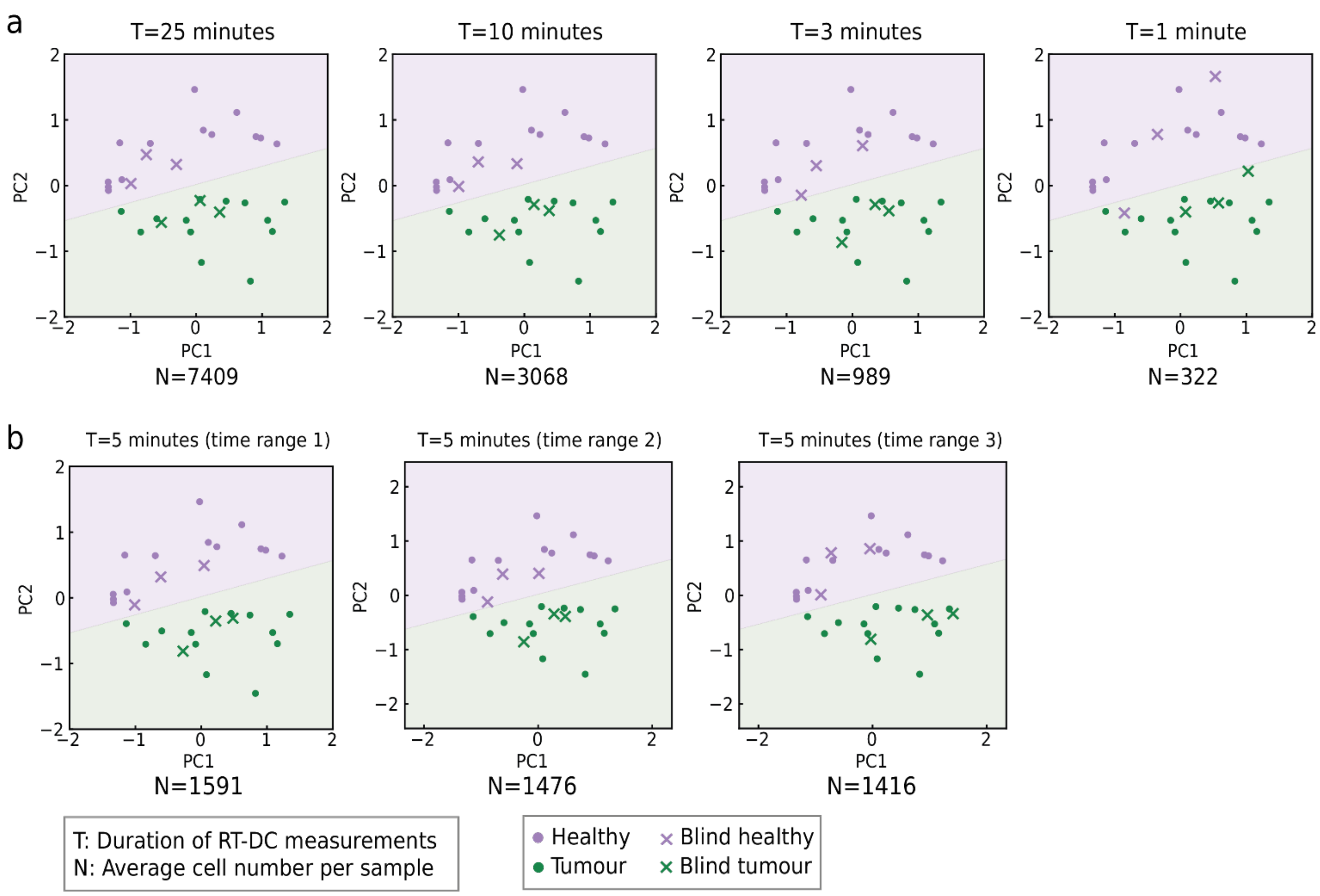


**Supplementary Fig. 9: Testing the performance of healthy vs tumour classification on reduced data from frozen human colon samples. a,** Logistic regression was performed on the PCA (shown by the linear divide in the PCA plot) and used to predict the classification of six blind samples (shown as crosses). From left to right, the amount of data used for classification decreases (the plots correspond to experiment durations of T = 25, 10, 3 and 1 minute). The average cell numbers analysed for a sample are shown below each plot (N). Classification was correct for experiments lasting 3 minutes or longer; data yielded by a one-minute experiment was insufficient for 100% correct classification of healthy vs tumour samples. **b,** PCA plots performed using approximately 1500 cells (or an equivalent of 5 minutes of measurement) extracted from different time points of longer RT-FDC measurements.

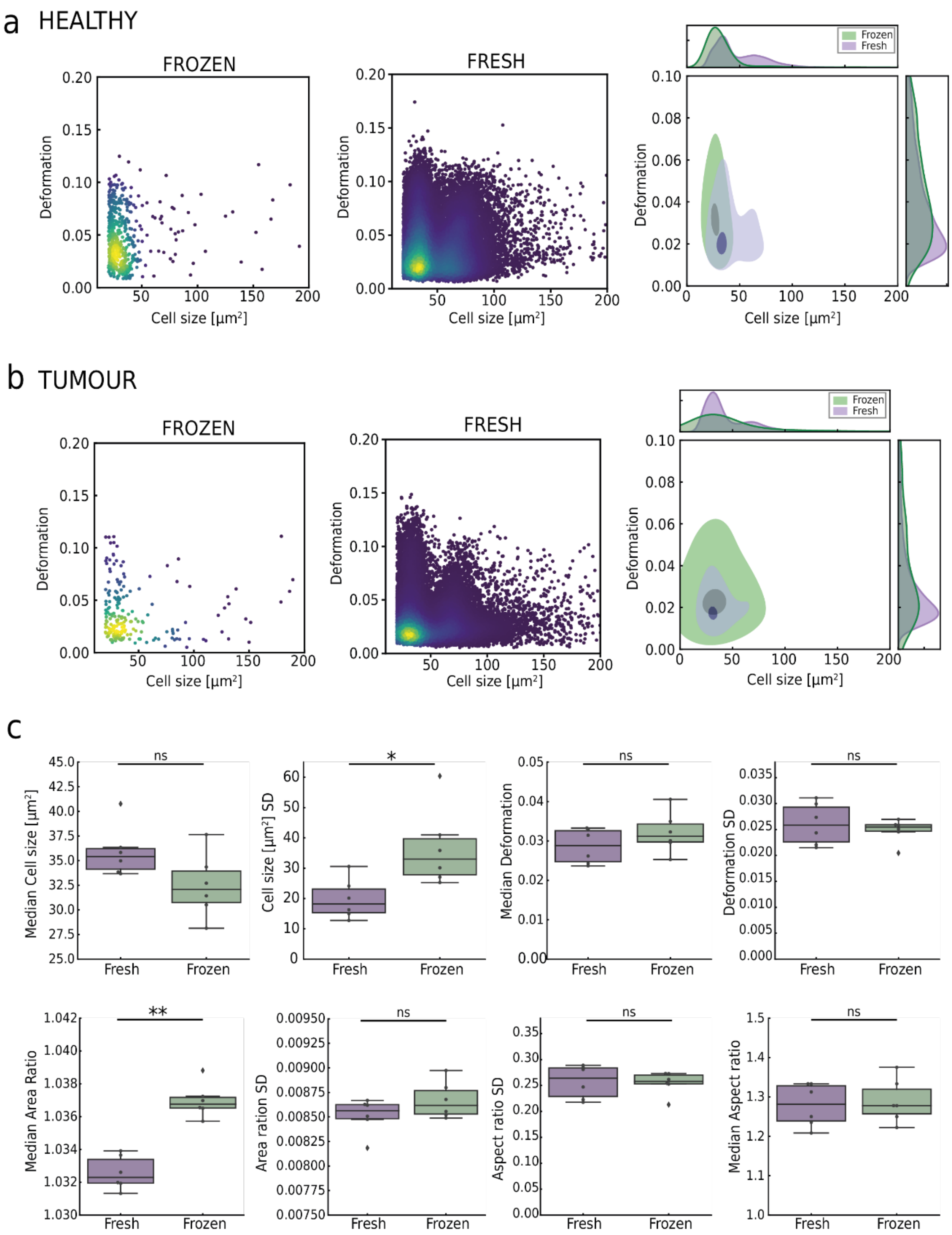


**Supplementary Fig. 10: Comparison of physical phenotype parameters of cells from frozen and fresh human biopsy samples.** Cell size vs deformation scatter plots of single cells extracted from either fresh (purple) or frozen (green) colon biopsy samples; **a,** healthy sample; **b,** tumour sample. The contour plots on the right correspond to the scatter plots on the left; the histograms show the distributions of cell size and deformation. **c,** Medians and standard deviations of cell size, deformation, area ratio and aspect ratio of fresh and frozen samples. Statistical comparisons were performed using Student's t-test, *p*-values are represented by * $p < 0.05$, ** $p < 0.01$, ns: non-significant.
